# IQOS product use in adults who smoke cigarettes daily does not change respiratory inflammatory markers

**DOI:** 10.1101/2025.11.17.25339937

**Authors:** Jennifer L. Thies, Hannah J. Appleseth, Naosuke Yamaguchi, Pria G. Bose, Adam M. Speen, Natalia Peraza, Catalina Cobos-Uribe, Alexia A. Perryman, Alayna P. Tackett, Meghan E. Rebuli

## Abstract

**Introduction:** The use and availability of heated tobacco products has increased globally. However, the health effects of IQOS product use remain contradictory, and the impact of IQOS product use on respiratory health and immune biomarkers is lacking. Industry sponsored studies suggest IQOS products reduce exposure to harmful chemicals compared to conventional cigarettes and show attenuated inflammatory responses compared to cigarette smoke, yet independent research studies indicate exposure to IQOS is still associated with negative health outcomes similar to conventional cigarettes. Clinical data is needed to understand the respiratory health impacts of IQOS use in people who currently smoke cigarettes.

**Methods:** Adults located in Los Angeles, California were recruited for the Adult IQOS Respiratory Study (AIRS, N = 32; 21-65 years). Nasal epithelial lining fluid (NELF) and saliva collected from adults who were willing to incorporate IQOS into their daily habits, were analyzed by multiplex enzyme linked immunosorbent assays for 29 different inflammatory markers or cotinine, respectively. Repeated measures ANOVA were used to identify changes in immune markers across the weekly visits. Linear mixed modelling was used to identify changes in participant cigarette or IQOS HeetStick use over the study period. Day was added as a fixed effect.

**Results:** Between-participants variability in IQOS or cigarette usage was identified (p < 0.0001). IQOS and cigarette use did not significantly change across time and remained stable over the course of the 4-week study period. No significant changes were identified in soluble protein mediators across each week. Cotinine concentrations remained consistent, even when disaggregated by sex and visit.

**Conclusions:** All together, these data highlight that dual product use may be more likely in individuals who smoke cigarettes, particularly over the first month of incorporation, rather than completely switching to IQOS products when provided. Furthermore, incorporating IQOS did not significantly change nicotine exposure or nasal inflammatory biomarkers, suggesting, at least with nicotine exposure and nasal inflammation, that addition of heated tobacco products may not substantially reduce harm.

**What is already known on this topic:** Heated tobacco products are marketed as less harmful and smoke-free alternative products compared to conventional cigarettes; however, lack of respiratory data and contradictory reports suggest they may still pose risks and use may contribute to consequential health impacts. Short-term and industry sponsored studies are available, but limited clinical data exists on the respiratory and immunological impacts of IQOS product use.

**What this study adds:** In the follow repeated measures study, our findings provide clinical evidence there are no significant differences in nasal immune biomarkers in individuals that smoke, who incorporate IQOS into their lifestyles. While each participant varied in their IQOS and cigarette product use, no significant trends were observed over the four-week sampling period in overall IQOS or cigarette use. Additionally, when participants were provided with the IQOS option, they were more inclined to dual use both products rather than switch from cigarettes to only IQOS use in the 4 week period studied.

**How this study might affect research, practice or policy:** The following clinical study suggests there is an increased likelihood of dual use among individuals who are provided alternative tobacco products. Additional studies should consider evaluating health differences, including immunological markers, between individuals who dual use IQOS and other tobacco products, exclusively use IQOS, people who completely switch their product use and people who quit altogether to establish whether IQOS use reduces harmful exposures resulting in differential inflammatory responses when compared to cigarette use.

## Introduction

Conventional cigarettes (CC) have known consequences and risks of use and exposure including lung inflammation, impaired lung immunity (1), structural damage caused by genetic changes (2) and increased risk of respiratory diseases including chronic obstructive pulmonary disease (COPD) (3). Introduction of purported safer alternatives to the tobacco market has been progressively rising, including heated tobacco products (HTPs). HTPs, which were presented to the tobacco industry in 2014, are alternative cigarette products that avoid combustion by heating included tobacco below temperatures of CC (4). This allows for nicotine delivery without the carcinogens and chemicals produced by burned tobacco – creating a possibly less harmful product compared to CC.

IQOS, a popular HTP available globally in over 60 countries, was authorized by FDA for sale in the US as a modified risk tobacco product (MRTP) in 2020 (5). This authorization allowed Philip Morris International (PMI), to claim their products reduce exposure risk to dangerous chemicals and products compared to cigarette smoke (CS). Studies have indicated attenuated inflammatory responses in epithelium cultures (6) and slowed progression of cardiovascular and respiratory changes in mice (7) compared to CS. Human-based studies indicated lower exposure levels to harmful and potentially harmful chemicals and sustained reductions in exposure biomarkers (8). However, these studies were conducted by PMI, and the human-based study focused on reduced outcomes in individuals who smoke cigarettes and were transitioning to HTP use. These studies failed to evaluate inflammatory and lung-specific immunological components of HTP use. It is currently unclear whether dual CC and HTP use, such as in the process of switching products, alters or improves respiratory outcomes over sole CC use in humans. Prior studies have primarily evaluated the effectiveness of switching, without evaluating changes in respiratory biology (9,10).

Industry independent research indicates HTPs still pose risks in healthy subjects and animal models, including induction of oxidative stress events and transcriptional changes in bronchial epithelial cells and alveolar mucosa models (11), increased proinflammatory biomarkers and compromised immune cell function in mouse lung (12) and increased cytotoxicity and inflammatory biomarkers compared to CS and e-cigarettes *in vitro* (13,14). These risks should especially be acknowledged as these products are not only being adopted by individuals who currently smoke cigarettes attempting to switch to a lower risk product or quit smoking. Studies have found increased adoption of HTPs among individuals with no history of smoking and those who previously smoked, including youth and young adults, (15,16). Altogether this raises the important question: what is the respiratory impact of IQOS product use? To answer this, we aimed to understand whether incorporation of IQOS products resulted in reduced CC use and how IQOS product use impacted respiratory inflammation. The following study reports on the respiratory impact of IQOS use, from adults who smoke cigarettes daily, but were willing to try incorporating IQOS use for 30 days. Participants were not required to eliminate use of cigarettes during the study. We evaluated respiratory immune mediators and cotinine concentrations, from nasal epithelial lining fluid (NELF) and saliva samples, respectively, over a 4-week period.

## Methods

### Primary Study Cohort Overview

Adults between 21 and 65 years of age were recruited for study participation in greater Los Angeles area of Southern California, USA between January – December 2021. Inclusion and exclusion criteria have been previously described (17), but briefly, participants were required to have a) ownership of a smartphone and willingness to download applications; b) currently daily smoking (more than 5 cigarettes/day for the past 6 months) c) age 21-65; and d) fluency in English e) never having used IQOS previously and f) willingness to try to IQOS use for the next 30 days. While the overall goal of the study was to evaluate potential product switching behavior, from cigarettes to IQOS, participants were not required to stop cigarette smoking during the study. Participants were excluded if they a) self-reported diagnosis of lung disease including cystic fibrosis, asthma, or chronic obstructive pulmonary disease b) had unstable or significant psychiatric conditions c) a history of cardiac event or distress within the past 3-months or d) currently pregnant or breastfeeding or planning to become pregnant or e) could not have used other tobacco products or marijuana more than 5 days of the past 30 (17). This protocol was reviewed and approved by the University of North Carolina – Chapel Hill, The Ohio State University, and The University of Southern California, Institutional Review Boards (IRB# 21-1078, 2023H0116, and UP-20-01209).

### Sample Collection and Processing

Nasal epithelial lining fluid (NELF) and saliva samples were collected as previously described (18,19) and stored at home in participant freezers until scheduled return pickup service was completed. Samples were collected 4 times, occurring once weekly during each study check-in visit conducted virtually and over videoconferencing. Samples were processed as previously described (19,20). As a nasal sampling strip was collected from both participant nares at each of the weekly samplings, a strip was randomly selected from either nostril for each participant at each visit to be used for downstream analyses. Prior studies suggested that there is limited variability from nare to nare in individuals (19). NELF was eluted as described previously with 1% BSA with 0.05% Triton X-100 in Dulbecco’s phosphate-buffered saline (DPBS) and centrifugation. The resulting NELF eluate was stored at −20°C until analysis. Saliva samples were collected via manufacturer directions procedures (Saliva Collection Aid, Salimetrics, State College, PA). Participants collected passive drool into a collection tube. Saliva was stored at –20°C until analysis.

### Salivary Cotinine Measurement

Cotinine, a nicotine metabolite, was used to evaluate exposure to nicotine-containing products. Cotinine was measured in saliva samples from each participant via competitive ELISA per manufacturer instructions (Salivary Cotinine ELISA Kit, Salimetrics, Carlsbad, CA (Cat No: 1-2002). For samples were diluted by a factor of 10, per manufacturer’s recommendation. For samples below the limit of detection for the assay, a value of zero was assigned.

### Protein extraction and measurement of cytokine levels

Proteins were analyzed with multiplex ELISA kit according to manufacturer instructions (V-PLEX Human Cytokine 30-plex, Mesoscale Discovery, Rockville, MD). The final analysis yielded measurements across 29 total cytokines, with concentration values in pg/mL for each sample collected. Concentration values were adjusted for differences in processing and dilution volumes. Dilutions were conducted based on manufacturer recommended minimum dilutions for each kit.

### Data Analysis

All analyses were conducted using R, v. 4.3.1 using base R unless otherwise noted (21). Linear-mixed models were conducted using *lme4 (22), lmerTest (23)*, and *zoo (24)*. Random intercepts for participants (record_ID) were used to account for within-subject variability for repeated measures across the study. Day was added as a fixed effect cofactor for random intercepts model. Sex was added as both a cofactor and interaction term in sensitivity analyses but did not improve model fit or display any significant interaction, so it was not included in the final model. Missingness total use entry data was handled by the last observation carried forward (na.locf) multiple imputation method under *imputeTS* (25). Plots were created with *consort (26), ggplot2 (27)* and *sjPlot (28)*. Participants that did not have a complete set of cytokine data from all four visits were removed from the final data set (*Supplemental Figure 1*). Missingness based on values below the assay limit of detection for cytokine data was handled by imputation. The value imputed was determined by taking the minimum value for each analyte dataset and dividing by 2. Normality was evaluated using a Shapiro-Wilk test. A non-parametric Kruskal-Wallis, followed by Dunn’s test for multiple comparisons, was used for statistical comparisons between groups.

## Results

### Demographics

A total of 32 individuals were recruited for the Adult IQOS Respiratory Study (AIRS) (17). 31 were enrolled and 24 completed the study. The average age of the cohort was 49.58 years, with participants ranging from 26 – 63 years of age. 12 individuals were female (50%). Most participants identified as white (39.1%) or black (43.5%) and non-Hispanic (83.3%). Nasal immune mediator analyses were conducted on 19 subjects (79.15%) that had complete cytokine data across all study visits. Of the 19, 57.9% were female (n=11) and 42.1% (n=8) were male.

### Participant-level variability in product use

To understand whether participants differed in their average use for either IQOS or CC, we examined random intercept only models (*Supplemental Table 2*). The average IQOS use across participants was 6.03 heat sticks per day. Participants differed significantly in their average IQOS use (var = 24.57; *p* < 0.0001). 41.6% (n=10) participants exhibited higher average IQOS use, while 58.3% (n=14) displayed lower than average use over the course of the study (*Figure 1A*). Similarly, for CC use, participants differed significantly in their average use (var = 17.05; *p* < 0.0001). The average CC use for participants was 3.34 cigarettes per day. 37.5% (n=9) participants exhibited higher than average CC use, while 62.5% (n=15) demonstrated lower than average use across the study (*Figure 1B*). These data indicate participants vary greatly in their average use habits for both products.

**Figure 1.**
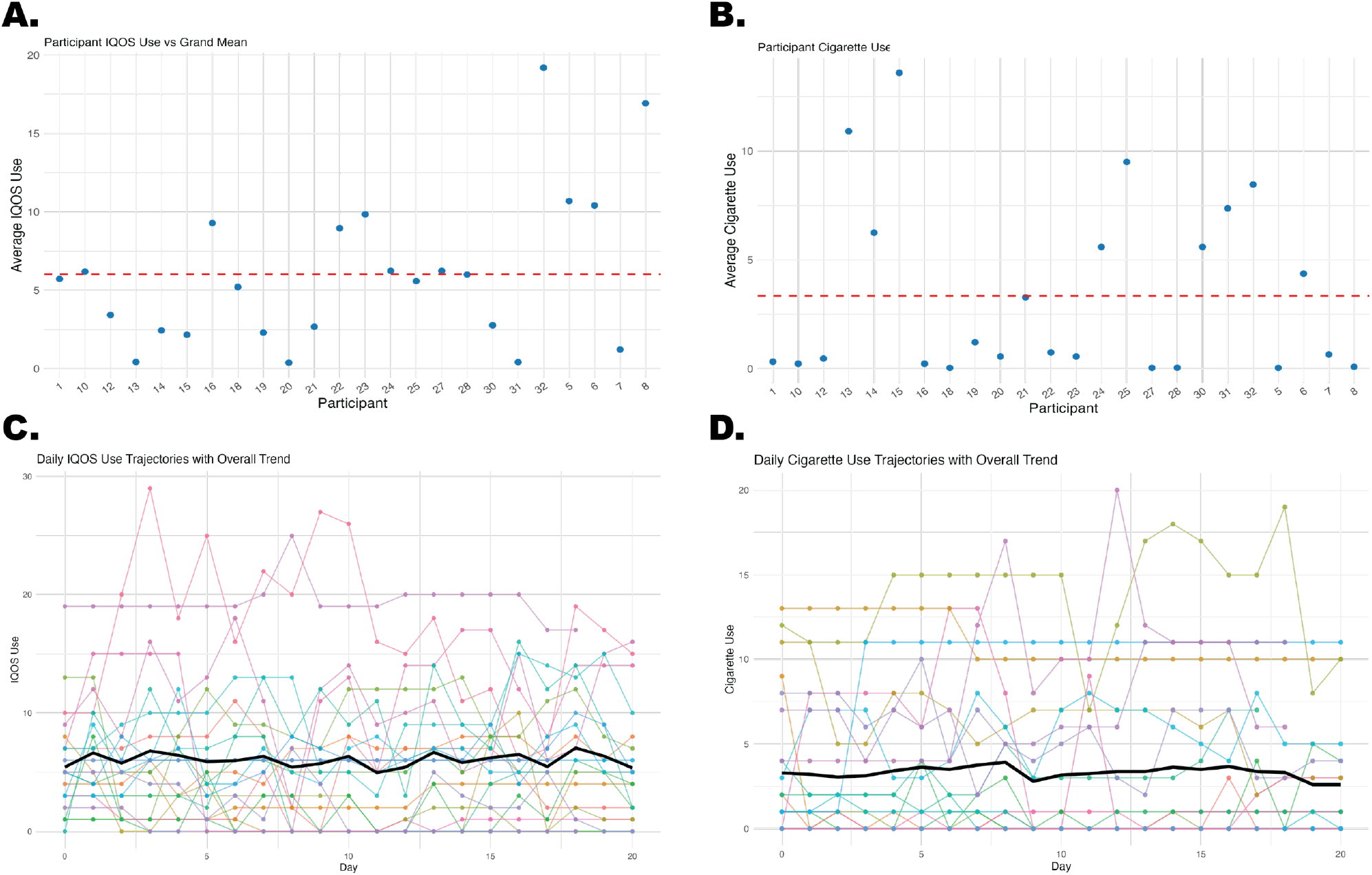
Participant IQOS and Cigarette use does not change over 4 weeks after initial IQOS introduction. (A) Estimated random intercepts for participants IQOS use. Points represent individual participant deviations from the overall mean (6.03). Red dotted line signifies the grand IQOS mean. (B) Estimated random intercepts for participant cigarette use. Points represent participant deviations from the overall mean (3.43). Red dotted line signifies the grand cigarette mean. (C) Daily IQOS use by participant. Each participant is denoted by a different colored line. Black solid line indicates daily average across participants. (D) Daily cigarette use by participant. Each participant is denoted by a different colored line. Black solid line indicates daily average across participants.

### IQOS and CC use do not differ over time

We were interested in whether time influenced average use patterns. With day as a fixed effect in our LMM, no significant difference was identified for IQOS (*p* = 0.91) or CC (*p* = 0.79) use (*Figure 1C and D*). When added as a fixed-effect, day does not significantly improve either model (*Supplemental Table 3*). Therefore, these data indicate average IQOS and CC product use remain stable throughout the study period, with no indications of overall increase or decrease use with time. This suggests that IQOS introduction did not reduce CC use in our cohort.

### Immune biomarkers remain stable over time

Limited respiratory and immunological data exist with IQOS introduction and use. NELF samples were collected once weekly to evaluate whether immune mediators changed over time with the addition of IQOS in individuals who smoke cigarettes. For all 29 measured analytes, no differences were identified across time (*p* > 0.05) (*Figure 2A-D*), suggesting the introduction and use of the IQOS product does not alter respiratory immune biomarkers compared to prior sole CC use.

**Figure 2.**
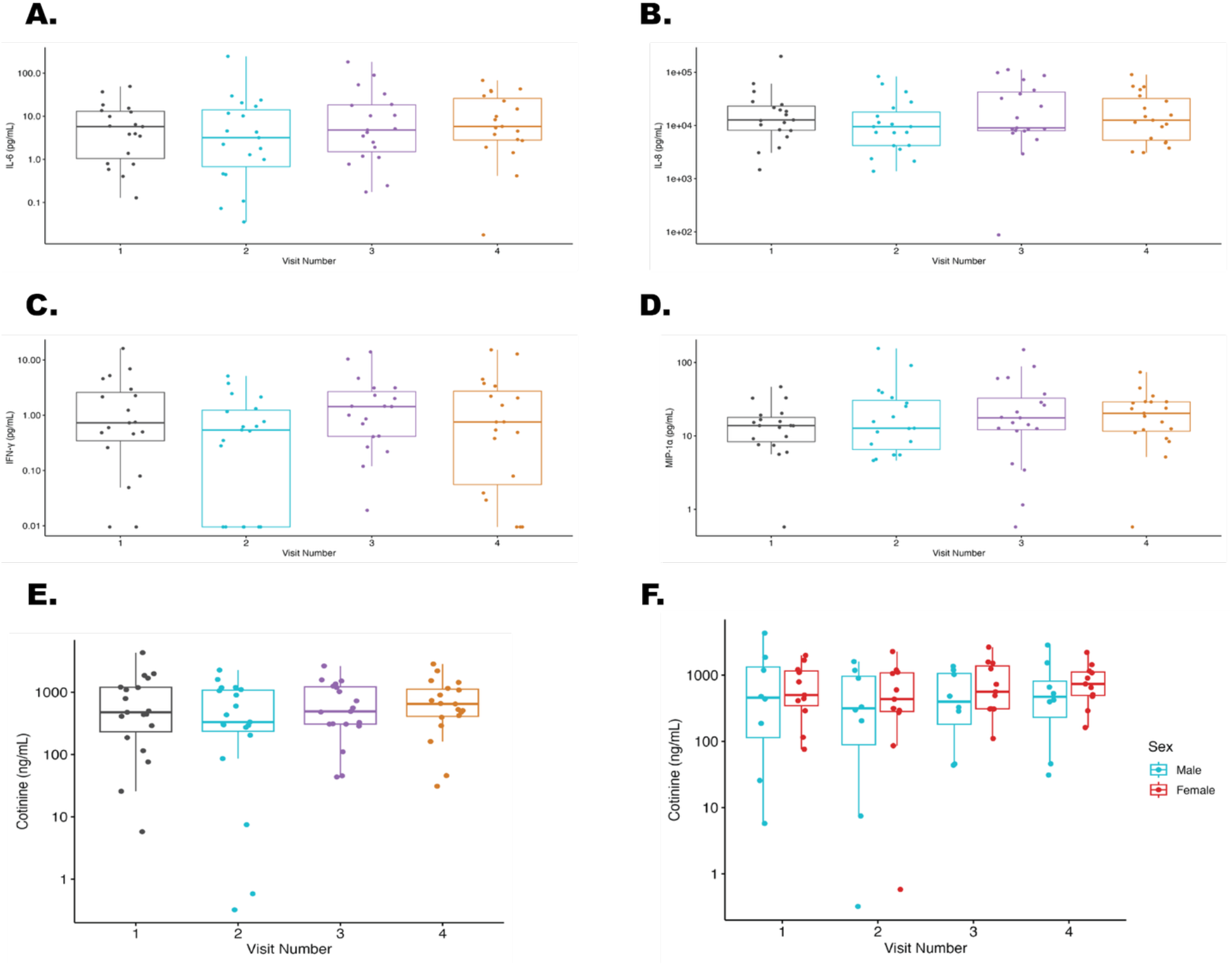
Soluble protein biomarkers and salivary cotinine remain unchanged across visits. (A) IL-6 (B) IL-8 (C) IFN-y and (D) MIP-1a do not change over time with IQOS addition and continued CC use. (E) Average aggregate salivary cotinine measured at each visit (F) Average salivary cotinine at each visit, separated by sex.

### No variation in cotinine concentration

Cotinine, a derivative of nicotine, was measured from saliva. Average cotinine for the group was 786.60 ng/mL. By sex, females had a cotinine average of 808.73 and males an average of 756.18 ng/mL (p > 0.05). No differences in cotinine concentration were identified between visits or sexes (p > 0.05) (*Figure 2E, F*). These data indicate nicotine exposure does not vary in individuals who dual use IQOS products and CC.

## Discussion

IQOS are becoming more readily available particularly in the United States, however knowledge of their impact on respiratory health is limited. In this preliminary pilot study, we investigated nasal cytokines from adults who smoke cigarettes daily, who were willing to incorporate IQOS products into their daily habits for 30 days. Overall, we discerned no significant differences in cytokine signatures or use patterns over the sampling period. Both IQOS and CC use remained relatively stable across time between participants, regardless of baseline use patterns. Likewise, nicotine exposure remained unchanged over the study period. To evaluate whether participants adopted IQOS products, we used LMM to evaluate trends in participants IQOS and CC use. We found between-participant variation to be significant as each participant had different average IQOS and CC use throughout the study. However, no significant changes in IQOS or CC use were found with time in the study.

IQOS aerosols were previously found to induce proinflammatory immune mediators, creating a proinflammatory microenvironment leading to lung damage like CS compared to air controls (12). Further, long-term exposure to IQOS aerosols produced upregulation of cell death and inflammatory genes resulting in apoptosis induced emphysema in murine lung (29). Limited research exists on dual use of IQOS and CC, however, *in vitro* studies suggest increased physiological and metabolic damage to airway epithelium compared to either product alone (30). Previous studies have noted similar salivary cotinine levels between people who exclusively use HTPs and individuals who use CCs. Similarly, IQOS and CC were found to yield nearly identical nicotine content and freebase nicotine under similar puffing regimens in other studies (31,32).

These data suggest HTP use is accompanied with similar CC risks and nicotine consumption, increasing the likelihood of sustained or developing addiction, particularly among people who do not smoke adopting use of IQOS without prior nicotine use. The misleading nature of IQOS being marketed as “smokeless and smoke-free” cigarette alternatives increases consumer interest. Ghazi and colleagues reviewed available literature on the health impacts and toxicity of IQOS and compared independent researcher data to industry sponsored studies and noted discrepancies between both sources (33). Ultimately, they concluded IQOS may not always coincide with the reduced risk potential that is advertised compared to cigarettes.

In the only other prior CC to IQOS adoption study examining respiratory health endpoints, Bhat et al., (2024) showed switching from CC to IQOS did not result in significant improvements in lung inflammation or damage compared to CC smoke exposure in vivo (34). While our study does not do a side-by-side comparison, we observed that IQOS use did not result in any attenuation of inflammatory cytokines. Our results here corroborate similar findings in the field that indicate switching to or the addition of IQOS products does not improve health effects compared to sole CC use and are likely not as “reduced risk” as claimed. These results are important as individuals who are adding IQOS use likely will not experience improvements in respiratory immune profiles. Further, these results suggest that IQOS use likely induces similar respiratory immune alterations to CC use, therefore adoption by never users or former smokers should be discouraged.

## Limitations

Limitations in the present study do exist. Increasing the number of eligible participants would provide increased power to potentially detect differences in biomarkers and use data. Additionally, the length of our study could have contributed to the absence of differences, with a study of longer duration potentially allowing for more complete switching to IQOS or manifesting biological impact of adoption later in time. Future studies assessing long-term respiratory health impacts of IQOS and dual use are required to affirm these results. Limitations with data collection were also present. Remote collection presented some difficulty for several participants, particularly pertaining to the application and troubleshooting issues with the IQOS product, and adopting use of the new product. Finally, participants were not mandated to completely switch their CC use to IQOS. Further studies should consider looking at groups with exclusive parameters of product use, including, exclusive users, dual smokers and switching smokers, as these factors may potentially show differences in respiratory immune outcomes.

## Conclusion

In general, this study indicated IQOS use in conjunction with CC does not result in significant immunological biomarker changes over a 4-week period. Likewise, nicotine exposure remained stable across time, suggesting addition of IQOS does not alter nicotine dose. The data highlight a lack of complete switching to IQOS products in individuals who are current daily CC users, regardless of baseline CC usage, and leads to dual use of tobacco products. This study further demonstrates the need for more clinical-based research to gain a full understanding of the health effects of IQOS use across the globe. Further, reassessment of whether IQOS products are providing reduced risk compared to CC should be considered, especially for respiratory health outcomes and with dual product use.

## Data Availability

All data produced in the present study are available upon reasonable request to the authors

## Acknowledgements

We would like to thank the research participants for the contributions to this study.

## Figures

**Supplemental Figure 1.**
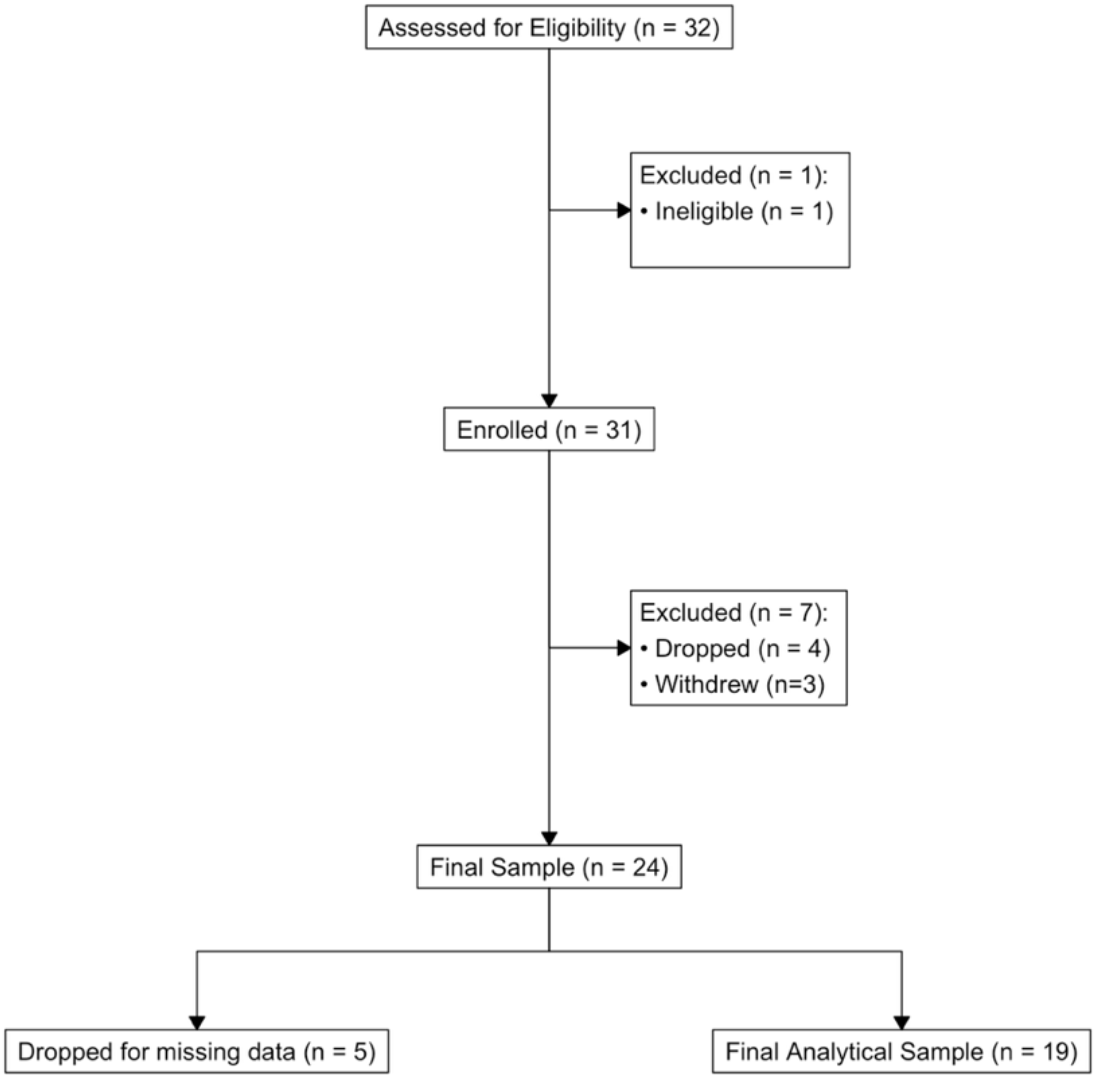
Consort Diagram

**Supplemental Table 2.**
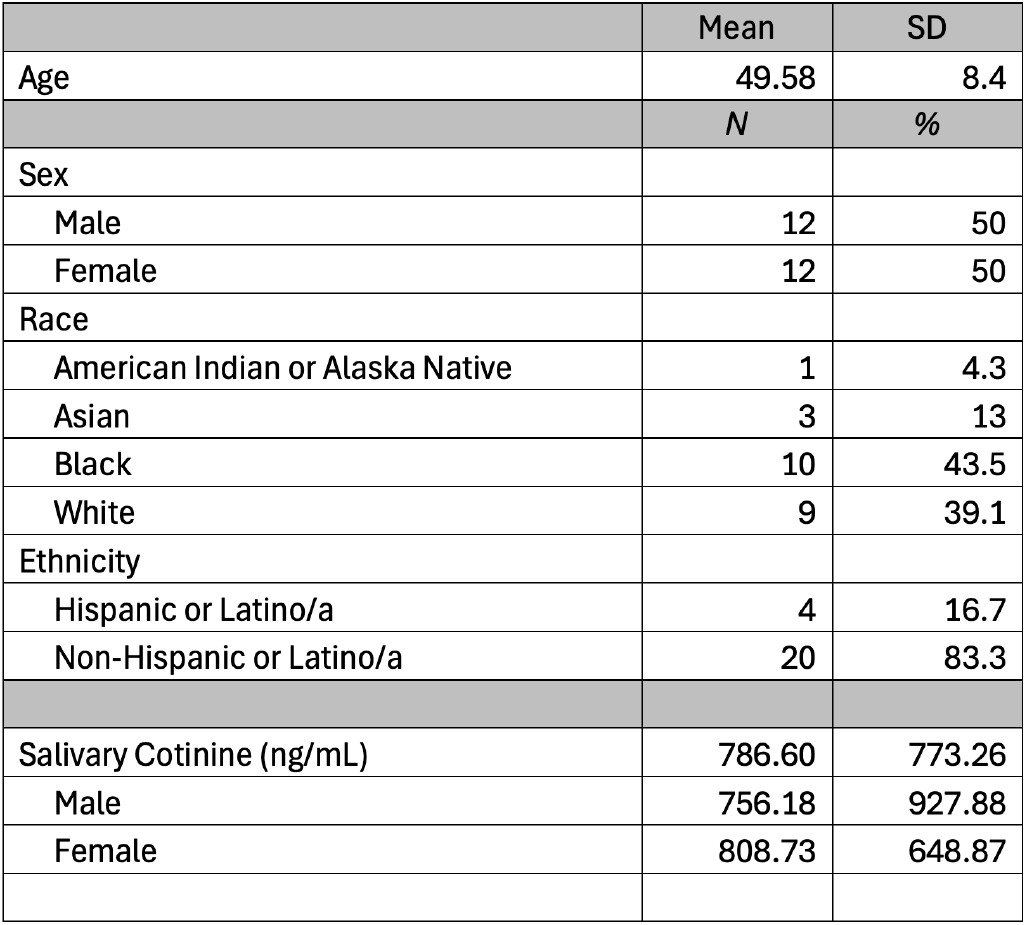
Demographics.

**Supplemental Table 2.**
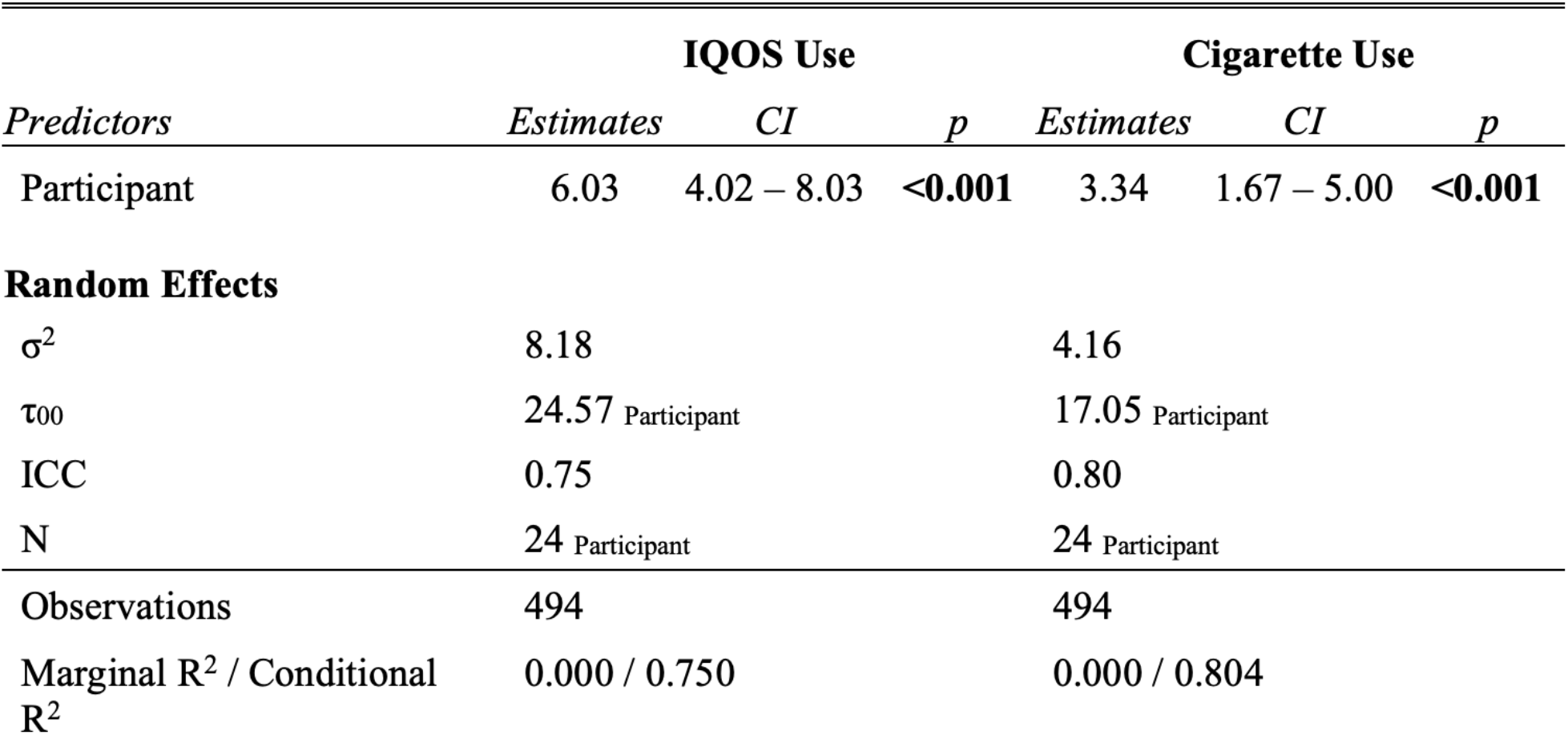
Unconditional Means for IQOS and Cigarette Total Use.

**Supplemental Table 3.**
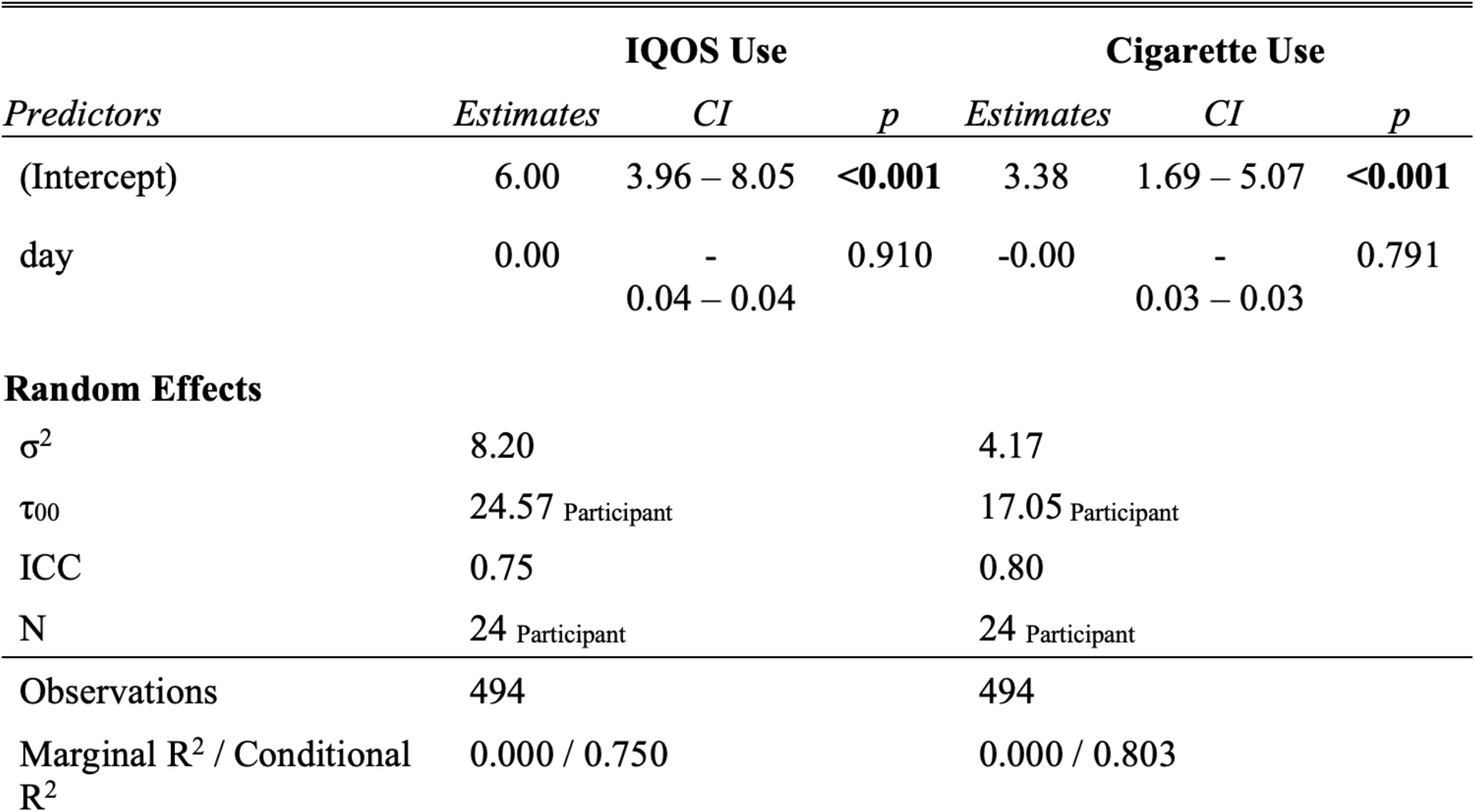
Random Intercepts for IQOS and Cigarette Use by Day.

