## Supplemental Figure 1 and Tables 1-3 for "IQOS product use in adults who smoke cigarettes daily does not change respiratory inflammatory markers"

Supplemental Figure 1 Consort Diagram

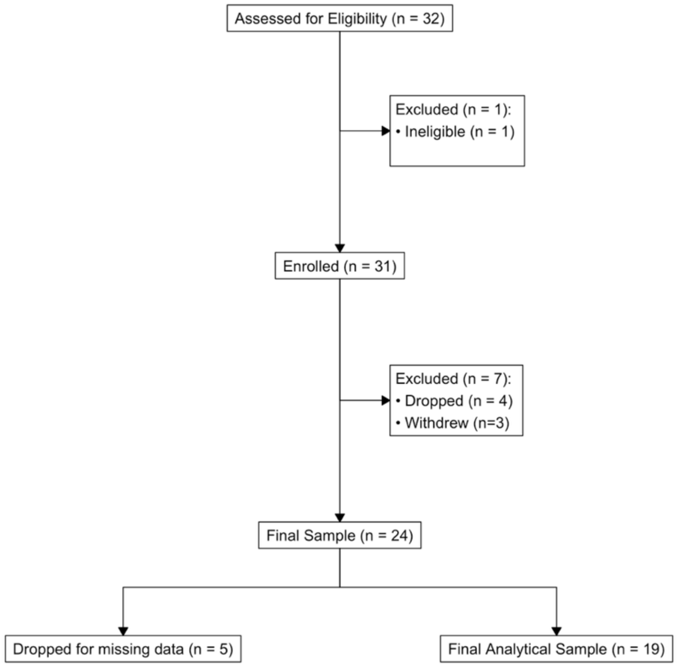

Supplemental Table 1 Study Demographics

| | Mean | SD |
|---|---|---|
| Age | 49.58 | 8.4 |
| | *N* | *%* |
| Sex | | |
| Male | 12 | 50 |
| Female | 12 | 50 |
| Race | | |
| American Indian or Alaska Native | 1 | 4.3 |
| Asian | 3 | 13 |
| Black | 10 | 43.5 |
| White | 9 | 39.1 |
| Ethnicity | | |
| Hispanic or Latino/a | 4 | 16.7 |
| Non-Hispanic or Latino/a | 20 | 83.3 |
| Salivary Cotinine (ng/mL) | 786.60 | 773.26 |
| Male | 756.18 | 927.88 |
| Female | 808.73 | 648.87 |

Supplemental Table 2 Unconditional Means for IQOS and Cigarette Total Use

| | IQOS Use | | | Cigarette Use | | |
|---|---|---|---|---|---|---|
| *Predictors* | *Estimates* | *CI* | *p* | *Estimates* | *CI* | *p* |
| Participant | 6.03 | 4.02 – 8.03 | **<0.001** | 3.34 | 1.67 – 5.00 | **<0.001** |
| **Random Effects** | | | | | | |
| $\sigma^2$ | 8.18 | | | 4.16 | | |
| $\tau_{00}$ | 24.57 Participant | | | 17.05 Participant | | |
| ICC | 0.75 | | | 0.80 | | |
| N | 24 Participant | | | 24 Participant | | |
| Observations | 494 | | | 494 | | |
| Marginal $R^2$ / Conditional $R^2$ | 0.000 / 0.750 | | | 0.000 / 0.804 | | |

Supplemental Table 3 Random Intercepts for IQOS and Cigarette Use by Day

| | IQOS Use | | | Cigarette Use | | |
|---|---|---|---|---|---|---|
| *Predictors* | *Estimates* | *CI* | *p* | *Estimates* | *CI* | *p* |
| (Intercept) | 6.00 | 3.96 – 8.05 | **<0.001** | 3.38 | 1.69 – 5.07 | **<0.001** |
| day | 0.00 | -0.04 – 0.04 | 0.910 | -0.00 | -0.03 – 0.03 | 0.791 |
| **Random Effects** | | | | | | |
| $\sigma^2$ | 8.20 | | | 4.17 | | |
| $\tau_{00}$ | 24.57 Participant | | | 17.05 Participant | | |
| ICC | 0.75 | | | 0.80 | | |
| N | 24 Participant | | | 24 Participant | | |
| Observations | 494 | | | 494 | | |
| Marginal $R^2$ / Conditional $R^2$ | 0.000 / 0.750 | | | 0.000 / 0.803 | | |
